# Sex-Specific Cord Blood DNA Methylation Signatures for Childhood ADHD Symptoms

**DOI:** 10.1101/2025.10.20.25338428

**Authors:** Markos Tesfaye, Solveig Løkhammer, Dinka Smajlagic, Anne-Kristin Stavrum, Kira D. Höffler, Christian M. Page, Jonelle D. Villar, Alexey Shadrin, Mona Bekkhus, Tetyana Zayats, Stephanie Le Hellard

## Abstract

**Background:** The underdiagnosis of Attention-Deficit/Hyperactivity Disorder (ADHD) in females, particularly those with the inattentive presentation, highlights a critical gap in clinical care. Although DNA methylation is often tissue-specific, accessible peripheral tissues like cord blood provide valuable insights into early-life risk and can serve as biomarkers. We aimed to identify sex-specific DNA methylation signatures in cord blood associated with childhood ADHD symptoms, which could illuminate early-life risk markers and inform improved detection strategies.

**Method:** We conducted sex-stratified epigenome-wide association study (EWAS) of cord blood DNA methylation in relation to ADHD symptoms in children (n=2,391; 48.1% females) from the Norwegian Mother, Father, and Child cohort study (MoBa). We tested for sex interaction effects and analyzed inattention and hyperactivity/impulsivity symptoms separately.

**Results:** We identified five differentially methylated CpG positions (DMPs) and 22 differentially methylated regions (DMRs). In females, two DMPs, cg13084029 (*ARRB1*) and cg15676735 (*ZBTB3*), and seven DMRs were associated with inattention symptoms. The identified DMPs exhibited significant sex interaction effects (adjusted *p*-value < 0.05). There was no overlap between the DMPs or DMRs identified in females and males, and the epigenetic signatures for inattention and hyperactivity/impulsivity were largely disparate. Several annotated genes (e.g., *ARRB1, PNPO, KDM5B*, *HOXA2,* and *HOXC4*) have recognized roles in neurotransmission and neurodevelopment.

**Conclusion:** Our findings demonstrate that the peripheral DNA methylation signatures at birth associated with later ADHD symptoms are distinct between females and males. This highlights the value of sex-stratified analyses and suggests that peripheral epigenetic profiles hold promise for the development of tools for early detection.

## Introduction

Attention Deficit Hyperactivity Disorder (ADHD) is a common neurodevelopmental disorder affecting 5 to 8% of children and adolescents globally,^1,2^ with symptoms persisting into adulthood in most affected individuals. While primarily characterized by pervasive inattentiveness or hyperactivity/impulsivity, it presents with a heterogeneous combination of symptoms, impacting clinical recognition and treatment.^3^ The higher prevalence in males than in females in childhood and adolescence^1,2^ tends to diminish in adulthood, partly due to significantly delayed recognition of females with predominant symptoms of inattention.^4,5^ Genetic and environmental factors contribute to ADHD risk,^6,7^ however, the molecular mechanisms underlying these sex differences in presentation and epidemiology are not fully understood.^8^

Epigenetic markers such as DNA methylation, which are influenced by genetic and environmental factors, offer a promising avenue to uncover early-life molecular signatures of ADHD.^9^ Considerable sex differences in brain DNA methylation profiles are documented across different stages of development,^10^ and these differences are also detectable in peripheral tissues, such as cord blood white cells at birth.^11^ While epigenome-wide association studies (EWASs) have begun identifying cord blood DNA methylation marks associated with ADHD,^12,13^ results across studies have been inconsistent.^14–17^ This heterogeneity may arise from differences in sample size, phenotype assessment methods, and a critical, yet often overlooked factor – failure to adequately account for sex differences in the analytical approach. To our knowledge, no prior EWAS has conducted a comprehensive, sex-stratified investigation of ADHD symptoms to identify female-specific and male-specific DNA methylation signatures.

It was recently demonstrated that meta-analyses accounting for sex can enhance epigenetic discovery when sex differences are present.^18^ Applying this approach to ADHD is particularly relevant given the reported sex differences in its epidemiology and clinical presentation.^1,2^ Therefore, we performed sex-stratified (male-only or female-only) and sex-differentiated (combining male-only and female-only through meta-analysis) analyses to identify DNA methylation signatures in cord blood associated with the total ADHD symptom score at the age of eight among children in the Norwegian Mother, Father and Child cohort study (MoBa). We also investigated DNA methylation marks associated with specific symptom domains of inattention and hyperactivity/impulsivity. Our goal was to identify early-life risk markers in cord blood that precede the onset of ADHD symptoms, providing insights into sex-specific pathways to ADHD and its subtypes, and contributing to the groundwork for future biomarker development that could improve early identification.

## Materials and Methods

### Cohort characteristics (MoBa)

The MoBa study is a nationwide longitudinal cohort with participants recruited between 1999 and 2008. On average 41% of the pregnant women during that period consented to participate. Overall, the MoBa includes approximately 114,500 children, 95,200 mothers and 75,200 fathers.^19^ The current study was based on version 12 of the quality-assured data files released for research in 2022. The establishment of MoBa and the initial data collection were approved by the Norwegian Data Protection Agency and the Regional Committees for Medical and Health Research Ethics. Participating mothers provided written informed consent. The MoBa is regulated by the Norwegian Health Registry Act. The current study was approved by the Regional Committees for Medical and Health Research Ethics (REK 2018/1898).

### DNA methylation data

This study was nested within the MoBa and included four sub-cohorts of children for whom DNA methylation was obtained in different batches. The sub-cohorts correspond to MoBa1 (met001), MoBa2 (met002), MoBa4 (met004), and MoBa8 (met008) sub-studies. Cord blood sample collection, phenotype, and covariate data collection were similar. (https://github.com/folkehelseinstituttet/mobagen/wiki/Methylation#documentation-valid-for-all-data-sets)

The MoBa1 sub-study initially comprised a random sample of children, including cases of asthma (n=1,128). DNA methylation typing was assessed in 2011 using Illumina Infinium HumanMethylation450 (450K) array (Illumina, San Diego, CA). Participants with complete data on ADHD symptoms and all covariates (n=646) were included in the current analysis. The MoBa2 sub-study consisted of a random sample (n=813) of children with and without asthma who had DNA methylation typed in 2013 using the 450K array.^11^ Among these, a subset (n=334) who had complete data on ADHD symptoms and covariates were included. The MoBa4 sub-study included a random sample of naturally conceived (n=983) and assisted reproductive technology conceived (n=962) children with DNA methylation data.^20^ Of these, 904 children with complete data on ADHD symptoms and covariates were included in the current study. The MoBa8 sub-study comprised a random sample of children selected for a diabetes and obesity study (n=1,238). From this cohort, 507 children had complete data on ADHD symptoms and covariates, and were included. DNA methylation profiling for MoBa4 and MoBa8 was performed using the Illumina Infinium HumanMethylationEPIC V1 array in 2019 and 2021, respectively (Illumina, San Diego, CA). The reasons for exclusion and numbers excluded for each sub-study are presented in a flow chart (Supplementary file: Figure S1).

### Phenotype and covariate data

#### ADHD symptoms

were assessed using the Parent/Teacher Rating Scale for Disruptive Behavior Disorders (RS-DBD) at the age of eight.^21^ RS-DBD has 18 items based on the DSM-IV ADHD symptoms. Nine of the items were related to the inattention cluster of symptoms, while the remaining nine items inquired about hyperactivity and impulsivity. Each item is rated on a 4-point Likert scale ranging from 1 to 4, with higher scores indicating greater symptom burden. The total ADHD symptom score was calculated from the responses to all 18 items, ranging from 18 to 72. The ‘*inattention*’ and ‘*hyperactivity/impulsivity*’ subscale scores were computed from the corresponding nine items and ranged from 9 to 36. The RS-DBD questionnaires were filled out by the mothers of children included in the MoBa.

#### Covariates

sex, birth weight in grams, and gestational age in weeks were recorded at birth and obtained from the Medical Birth Registry of Norway. We computed smoking scores from DNA methylation data.^22,23^ Similarly, estimated blood cell-type proportions were obtained by applying the Houseman method as described previously.^12,24^

### DNA methylation data

Details of the protocol for collecting and storing cord blood samples in the MoBa are described elsewhere.^25^ In brief, umbilical cord blood samples were collected at birth and stored at -80 degrees Celsius at the study Biobank.^25^ Bisulfite conversion was performed using the EZ-96 DNA Methylation kit (Zymo Research Corporation, Irvine, CA).^12^

Quality control and data processing were performed independently for each of the four sub-cohorts. The intensity (IDAT) files were processed using the *minfi* package to generate methylation level measure (beta values) for each cytosine-phosphate-guanine (CpG) site (i.e., the fraction of methylated sites per sample calculated as the ratio of methylated and unmethylated fluorescent signals in the mixture of blood cells) and detection *p*-values. Background subtraction was performed using out-of-band probes (ss-Noob). The Beta Mixture Quantile (BMIQ) method was applied to normalize for the differences in the Illumina probe designs.^26^ Control probes, non-autosomal probes, and CpGs with > 10% missing methylation data were removed. Samples with an average detection *p*-value across all probes < 0.05 and samples with a mismatch between the reported and methylation-derived sex were also removed.^12^ Detailed quality control steps are provided at: https://github.com/folkehelseinstituttet/mobagen/wiki/MethylationQC.

### Epigenome-wide associations

Three sets of EWASs were conducted, focusing on the total ADHD symptoms, the inattention domain, and the hyperactivity/impulsivity domain. All EWASs were performed as sex-stratified; therefore, they comprised data from only female or male children. For each EWAS, linear regression analysis was performed using the *limma* package to identify differentially methylated CpG positions (DMPs) associated with the respective ADHD symptom scores.^27^ The methylation beta values for each probe were regressed against the symptom scores (total ADHD symptom scores, inattention domain, or hyperactivity/impulsivity domain scores) in a model that included the following covariates: birth weight, gestational age at delivery, cord blood cell type proportions, smoking scores, two ancestry principal components (PCs) computed from methylation data,^28^ sample plate, five methylation PCs, and 30 control probe PCs. Quantile-quantile (QQ) plots and lambda values (λ) from each EWAS were examined for any inflation (Supplementary file: Figures S2 – S7). Probes mapped on the sex chromosomes (X and Y) were excluded from the analyses.

### Meta-analyses

Meta-analyses were performed for sex-stratified (male-only or female-only) and sex-differentiated (combination of all male-only and female-only summary statistics) models. The sex-stratified meta-analysis combined results from four EWASs, and the sex-differentiated meta-analysis included results from eight EWASs (four female-only and four male-only). In all meta-analyses, effect sizes and standard errors were analyzed using the *metagen* function of the *meta* R-package. The implementation employs a standard inverse variance meta-analysis, where the pooled effect sizes are calculated as the weighted average of the effect sizes in each EWAS. Probes with results from EWAS in at least two sub-studies were included in the meta-analyses. Based on the evidence presented on adequate control for false positive results,^29^ DMPs with *p*-value less than 9.0 × 10^-8^ on the fixed effects model were considered significant.

We applied the *comb-p* procedure to identify differentially methylated regions (DMRs).^30,31^ The specific parameters used were seed p-value□=□0.001 and maximum distance between probes of 750 base pairs. DMRs with a minimum of four probes were identified with a Sidak-adjusted p-value threshold of p < 0.05.^31^

### Functional regulation and gene set analyses

The identified DMPs and DMRs were annotated to genes using the ‘*annotatr*’ package.^32^ Annotations were obtained by selecting ’hg19_basicgenes’, which provided genes for DMPs and DMRs in the promoter, 5’UTR, 3’UTR, exon, intron, and 1kb to 5kb upstream regions. Enrichment and pathway analyses were investigated on genes annotated to the top 200 CpGs using the *Gometh* function and ‘EPIC array’ setting in *missMethyl*.^33^

We examined the regulation by genetic variants of the methylation status at the identified DMPs. This was achieved by exploring the DMPs for a list of known *cis*-acting methylation quantitative trait loci (*me*QTLs) in the human blood at birth.^34^

### Sex interaction effects, and overlap of DMPs and DMRs

Sex interaction effects were tested for the identified DMPs, and Bonferroni correction was applied to their *p*-values to minimize false discovery. First, a sex interaction (i.e., sex × ADHD score) term was added to the EWAS model as described above. The effect sizes and standard errors of the sex interaction term from each sub-study were meta-analyzed.

We examined the results for any overlap across identified DMPs and DMRs in females, males, and individuals with inattention-only and hyperactivity/impulsivity-only. Additionally, we further examined for any overlap of CpGs with suggestive *p*-values < 0.0001, and these overlaps are presented in a Venn diagram.

## Results

### Cohort characteristics

Complete data on ADHD symptoms, DNA methylation, and covariates were available for 2,391 (48.1% females) children. Table 1 provides the characteristics of the study participant children across the four sub-studies. The mean birth weights were higher for males (*p*-value < 0.05). On a non-parametric (Mann–Whitney) test, males had higher ADHD and inattention symptom scores than females in all four sub-studies (*p*-value < 0.05).

**Table 1:** Characteristics of MoBa study participant children included in epigenome-wide association study of ADHD symptoms.

|  | **MoBa1**<br>(n=646) |  | **MoBa2**<br>(n=334) |  | **MoBa4**<br>(n=904) |  | **MoBa8**<br>(n=507) |  |
| --- | --- | --- | --- | --- | --- | --- | --- | --- |
|  | Male | Female | Male | Female | Male | Female | Male | Female |
| Sample size (n) | 345 | 301 | 192 | 142 | 449 | 455 | 255 | 252 |
| Gestational age (weeks), mean (SD) | 40.0 (1.9) | 39.7 (1.8) | 39.8 (1.9) | 39.5 (1.7) | 39.7 (1.9) | 39.7 (2.5) | 40.0 (2.4) | 39.5 (3.1) |
| Birth weight (kgs), mean (SD) * | 3.7 (0.5) | 3.5 (0.5) | 3.7 (0.5) | 3.6 (0.5) | 3.6 (0.5) | 3.5 (0.5) | 3.7 (0.5) | 3.6 (0.6) |
| Cell proportion (%), mean (SD) |  |  |  |  |  |  |  |  |
| Granulocytes | 44.9 (9.5) | 47.0 (9.1) | 43.8 (9.3) | 46.5 (9.5) | 44.6 (9.9) | 47.3 (9.4) | 45.6 (9.5) | 47.2 (8.4) |
| B cells | 14.5 (3.8) | 14.9 (3.2) | 15.0 (3.8) | 14.2 (4.0) | 13.9 (3.7) | 13.2 (3.4) | 12.2 (3.9) | 11.7 (3.3) |
| Natural killer cells | 4.0 (3.4) | 4.2 (3.2) | 4.1 (3.4) | 4.1 (3.4) | 3.6 (3.1) | 3.9 (3.1) | 1.8 (2.7) | 1.9 (2.6) |
| CD4<sup>+</sup> T cells | 19.4 (5.1) | 19.3 (5.5) | 19.5 (5.3) | 18.2 (4.7) | 18.1 (5.1) | 17.7 (5.0) | 18.3 (5.8) | 17.8 (5.7) |
| CD8<sup>+</sup> T cells | 14.8 (5.0) | 13.5 (4.4) | 14.3 (4.5) | 13.7 (4.4) | 15.6 (4.8) | 13.6 (4.5) | 15.3 (4.3) | 14.4 (3.9) |
| Monocytes | 10.0 (2.4) | 9.5 (2.4) | 10.4 (2.3) | 10.7 (2.3) | 10.2 (2.3) | 10.0 (2.2) | 9.1 (2.5) | 9.4 (2.5) |
| Smoking scores, mean (SD) | 2.4 (2.1) | 2.5 (2.1) | 1.6 (2.0) | 1.7 (2.1) | -0.33 (2.0) | -0.07 (2.0) | 0.20 (1.9) | 0.39 (1.9) |
| ADHD score, median (IQR) * | 26 (8) | 24 (6) | 26 (9) | 23 (6) | 26 (8) | 24 (7) | 25 (8) | 23 (6) |
| Inattention score, median (IQR) * | 14 (5) | 12 (3) | 14 (5) | 12 (4) | 14 (5) | 13 (4) | 13 (4) | 13 (3) |
| Hyperactivity/impulsivity score, median (IQR) | 12 (5) | 11 (3) | 12 (5) | 11 (3.3) | 12 (5) | 11 (3) | 11 (4) | 11 (4) |
| (*) significant differences between males and females at *p*-value less than 0.05 |  |  |  |  |  |  |  |  |
| ADHD- attention deficit hyperactivity disorder; IQR – Interquartile range; SD – standard deviation |  |  |  |  |  |  |  |  |

### All ADHD symptoms and DNA methylation

In the total sample, ADHD symptoms were associated with two DMRs, chr17:6796906-6797709 and chr17:46018929-46019175 (Supplementary Tables: Table S1), but no DMPs reached epigenome-wide significance (Supplementary file: Figure S8). ADHD symptoms were associated with hypomethylation at cg16317998 (*FOXP4-AS1*) in males, and hypermethylation at cg15676735 (*ZBTB3*) in females (Figure 1 and Table 2).

**Figure 1.**
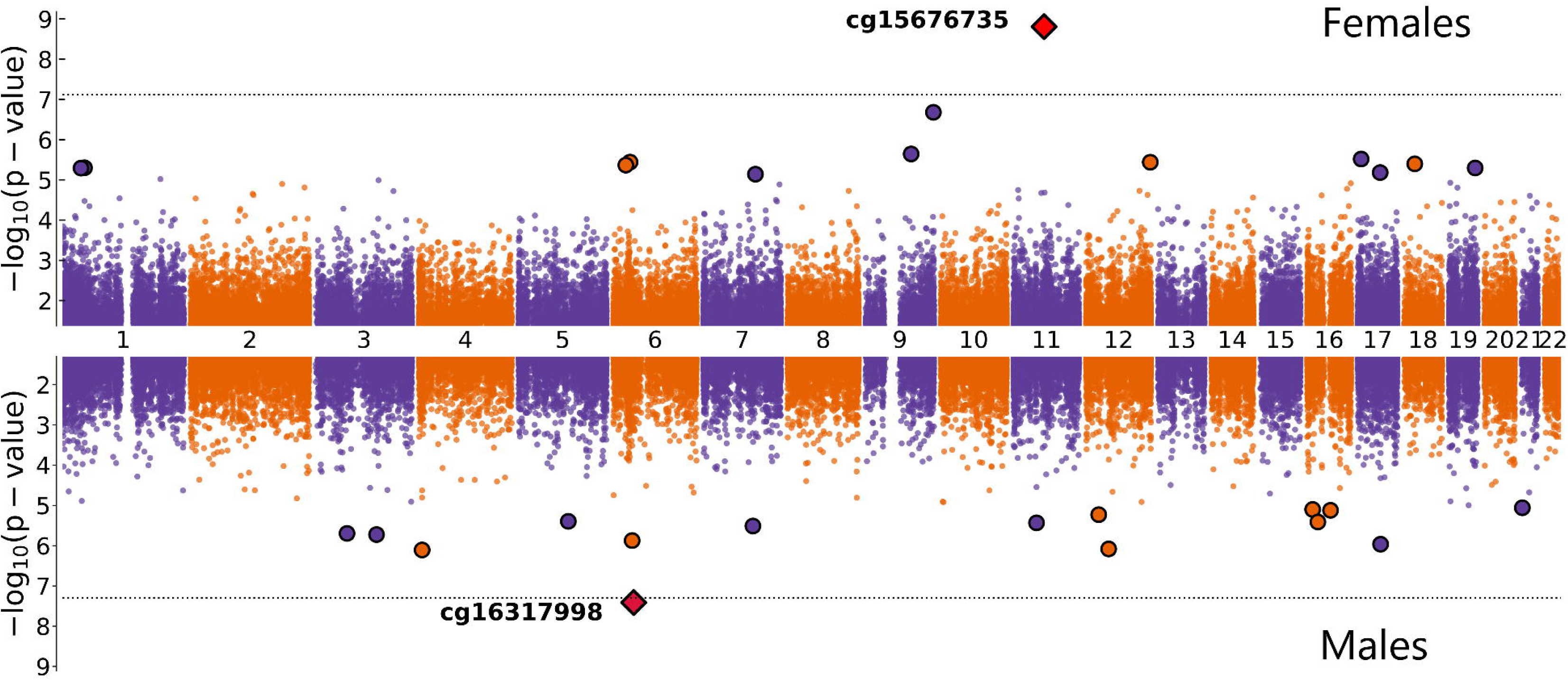
Miami plot showing differentially methylated CpG sites in cord blood associated with childhood ADHD symptoms in female and male participants of the MoBa study. Chromosomes are on the x-axis, -log10 of *p*-values on the y-axis, and the dotted horizontal line represents epigenome-wide significance (9.0 × 10^-8^).

**Table 2.** Cord blood DNA methylation loci associated with total ADHD, inattention, and hyperactivity/ impulsivity symptoms at the age of eight in children in the MoBa study.

|  |  | cg05928306 | cg13084029 | cg13426208 | cg15676735 | cg16317998 |
| --- | --- | --- | --- | --- | --- | --- |
| CHR: Position |  | 14:56351623 | 11:74994968 | 2:11795756 | 11:62521955 | 6:41454329 |
| Gene |  |  | *ARRB1* | *NTSR2* | *ZBTB3* | *FOXP4-AS1* |
| **Inattention symptoms** |  |  |  |  |  |  |
| Female | Effect | $-2.10 \times 10^{-4}$ | **$-2.89 \times 10^{-3}$** | $-9.22 \times 10^{-5}$ | **$1.70 \times 10^{-4}$** | $4.98 \times 10^{-4}$ |
| | SE | $2.21 \times 10^{-4}$ | **$5.24 \times 10^{-4}$** | $9.12 \times 10^{-5}$ | **$3.07 \times 10^{-5}$** | $3.43 \times 10^{-4}$ |
| | P-value | $3.42 \times 10^{-1}$ | **$3.42 \times 10^{-8}$** | $3.12 \times 10^{-1}$ | **$2.73 \times 10^{-8}$** | $1.46 \times 10^{-1}$ |
| Male | Effect | $-1.81 \times 10^{-5}$ | $3.25 \times 10^{-5}$ | $-1.07 \times 10^{-4}$ | $8.19 \times 10^{-6}$ | $-1.77 \times 10^{-3}$ |
| | SE | $5.91 \times 10^{-5}$ | $1.89 \times 10^{-4}$ | $7.92 \times 10^{-5}$ | $1.87 \times 10^{-5}$ | $4.73 \times 10^{-4}$ |
| | P-value | $7.60 \times 10^{-1}$ | $8.64 \times 10^{-1}$ | $1.78 \times 10^{-1}$ | $6.62 \times 10^{-1}$ | $1.88 \times 10^{-4*}$ |
| Total sample | Effect | $-3.09 \times 10^{-5}$ | $-3.06 \times 10^{-4}$ | $-1.00 \times 10^{-4}$ | $5.22 \times 10^{-5}$ | $-2.82 \times 10^{-4}$ |
| | SE | $5.71 \times 10^{-5}$ | $1.78 \times 10^{-4}$ | $5.98 \times 10^{-5}$ | $1.60 \times 10^{-5}$ | $2.78 \times 10^{-4}$ |
| | P-value | $5.88 \times 10^{-1}$ | $8.62 \times 10^{-2}$ | $9.29 \times 10^{-2}$ | $1.08 \times 10^{-3*}$ | $3.10 \times 10^{-1}$ |
| **Hyperactivity/impulsivity symptoms** |  |  |  |  |  |  |
| Female | Effect | **$-1.22 \times 10^{-3}$** | $-6.52 \times 10^{-4}$ | $-4.36 \times 10^{-4}$ | $1.60 \times 10^{-4}$ | $-5.31 \times 10^{-4}$ |
| | SE | **$2.23 \times 10^{-4}$** | $5.99 \times 10^{-4}$ | $1.03 \times 10^{-4}$ | $3.24 \times 10^{-5}$ | $3.57 \times 10^{-4}$ |
| | P-value | **$4.54 \times 10^{-8}$** | $2.76 \times 10^{-1}$ | $2.18 \times 10^{-5*}$ | $8.04 \times 10^{-7*}$ | $1.36 \times 10^{-1}$ |
| Male | Effect | $-1.01 \times 10^{-4}$ | $9.72 \times 10^{-5}$ | $-2.98 \times 10^{-4}$ | $8.84 \times 10^{-6}$ | **$-2.96 \times 10^{-3}$** |
| | SE | $6.20 \times 10^{-5}$ | $2.00 \times 10^{-4}$ | $8.25 \times 10^{-5}$ | $2.50 \times 10^{-5}$ | **$4.82 \times 10^{-4}$** |
| | P-value | $1.05 \times 10^{-1}$ | $6.27 \times 10^{-1}$ | $3.09 \times 10^{-4*}$ | $7.24 \times 10^{-1}$ | **$8.87 \times 10^{-10}$** |
| Total sample | Effect | $-1.81 \times 10^{-4}$ | $2.22 \times 10^{-5}$ | **$-3.52 \times 10^{-4}$** | $6.53 \times 10^{-5}$ | $-1.39 \times 10^{-3}$ |
| | SE | $5.98 \times 10^{-5}$ | $1.90 \times 10^{-4}$ | **$6.43 \times 10^{-5}$** | $1.98 \times 10^{-5}$ | $2.87 \times 10^{-4}$ |
| | P-value | $2.46 \times 10^{-3*}$ | $9.07 \times 10^{-1}$ | **$4.46 \times 10^{-8}$** | $9.85 \times 10^{-4*}$ | $1.29 \times 10^{-6*}$ |
| **ADHD symptoms** |  |  |  |  |  |  |
| Female | Effect | $-5.08 \times 10^{-4}$ | $-1.25 \times 10^{-3}$ | $-1.57 \times 10^{-4}$ | **$1.07 \times 10^{-4}$** | $9.84 \times 10^{-7}$ |
| | SE | $1.35 \times 10^{-4}$ | $3.38 \times 10^{-4}$ | $5.47 \times 10^{-5}$ | **$1.78 \times 10^{-5}$** | $2.16 \times 10^{-4}$ |
| | P-value | $1.74 \times 10^{-4*}$ | $2.19 \times 10^{-4*}$ | $4.21 \times 10^{-3*}$ | **$1.82 \times 10^{-9}$** | $9.96 \times 10^{-1}$ |
| Male | Effect | $-3.58 \times 10^{-5}$ | $3.79 \times 10^{-5}$ | $-1.24 \times 10^{-4}$ | $6.66 \times 10^{-6}$ | **$-1.47 \times 10^{-3}$** |
| | SE | $3.39 \times 10^{-5}$ | $1.12 \times 10^{-4}$ | $4.53 \times 10^{-5}$ | $1.42 \times 10^{-5}$ | **$2.67 \times 10^{-4}$** |
| | P-value | $2.91 \times 10^{-1}$ | $7.34 \times 10^{-1}$ | $6.08 \times 10^{-3*}$ | $6.38 \times 10^{-1}$ | **$3.89 \times 10^{-8}$** |
| Total sample | Effect | $-6.38 \times 10^{-5}$ | $-8.91 \times 10^{-5}$ | $-1.37 \times 10^{-4}$ | $4.55 \times 10^{-5}$ | $-5.80 \times 10^{-4}$ |
| | SE | $3.29 \times 10^{-5}$ | $1.06 \times 10^{-4}$ | $3.49 \times 10^{-5}$ | $1.11 \times 10^{-5}$ | $1.68 \times 10^{-4}$ |
| | P-value | $5.27 \times 10^{-2}$ | $4.01 \times 10^{-1}$ | $8.21 \times 10^{-5*}$ | $4.00 \times 10^{-5*}$ | $5.56 \times 10^{-4*}$ |
| Epigenome-wide significant associations are written in **bold**; (*) nominally significant p-values |  |  |  |  |  |  |
| ADHD – attention deficit hyperactivity disorder; CHR – chromosome; SE – standard error |  |  |  |  |  |  |

ADHD symptoms were associated with four DMRs, including chr17:46018731-46019018 (*PNPO*) in males, but with no DMRs in the total sample. In females, ADHD symptoms were associated with six DMRs, such as chr1:202778575-202778870 (*KDM5B*). (Supplementary Tables: Tables S2 & S3). *KDM5B* encodes a histone demethylase, an enzyme involved in epigenetic regulation of genes as well as plays a key role in neurogenesis.^35^

### Inattention symptoms and DNA methylation

Inattention symptoms were associated with five DMRs (e.g., chr11:6291339-6292616; *CCKBR*) in the total sample (Supplementary Tables: Tables S4), however, no DMPs were identified (Supplementary file: Figure S9). No DMPs or DMRs were identified for inattention symptoms in males (Supplementary file: Figure S10). In females, inattention symptoms were associated with two DMPs and seven DMRs (Supplementary Tables: Table S5). The two DMPs showed hypomethylation at cg13084029 (*ARRB1*) and hypermethylation at cg15676735 (*ZBTB3*) (Table 2, Supplementary file: Figure S11).

### Hyperactivity/impulsivity symptoms and DNA methylation

Hypomethylation at cg16317998 (in males), cg05928306 (in females), and cg13426208 (in sex-differentiated) meta-analyses was associated with hyperactivity/impulsivity symptoms (Table 2, Supplementary file: Figures S12-14). Four and six DMRs were associated with hyperactivity/impulsivity symptoms in males and females, respectively (Supplementary Tables: Tables S6 & S7). The DMR chr17:6796745-6797120 (*ALOX12P2*) was associated with both inattention and hyperactivity/impulsivity symptoms in females (Supplementary file: Tables S5 & S7). Two DMRs were annotated to functionally related *HOX* genes: chr7:27143235-27143586 (*HOXA2*) in males and chr12:54446289-54446538 (*HOXC4*) in females, both associated with hyperactivity/impulsivity symptoms.

### Functional regulation and gene set analyses

DMPs and DMRs located in the promoter, 5’UTR, 3’UTR, exon, intron, or within 1kb to 5kb upstream regions were annotated to the nearby genes (Table 2, Supplementary Tables S1-S7). None of the pathways in the gene set analyses for genes annotated to the top 200 CpGs reached statistical significance (false discovery rate > 0.05). None of the identified DMPs had known *cis*-acting variants in blood at birth listed in the *me*QTL database.

### Sex interaction effects, and overlap of DMPs and DMRs

Our analyses revealed that the association between cord blood DNA methylation and ADHD symptoms is predominantly sex specific. This is underscored by the finding that all DMPs identified in females exhibited significant sex interaction effects, confirming their association is specific or significantly stronger in females (Supplementary Tables: Table S8). Furthermore, the DMP identified in males was not nominally significant in females (Table 2).

To identify overlapping DNA methylation signatures between males and females, we examined a larger number of CpGs with *p*-values less than 0.0001 (Figure 2). None of the 406 CpGs overlapped between females and males. Furthermore, only nine of the 406 CpGs overlapped between inattention and hyperactivity/impulsivity (Figure 2). This pattern of non-overlap was also observed in DMR analyses, which identified distinct sets of DMRs in females and males (Supplementary Tables: S2-S3 & S6-S7). Overall, the cord blood DNA methylation profiles associated with childhood total ADHD, inattention-only, and hyperactivity/impulsivity-only symptoms showed little overlap between males and females.

**Figure 2.**
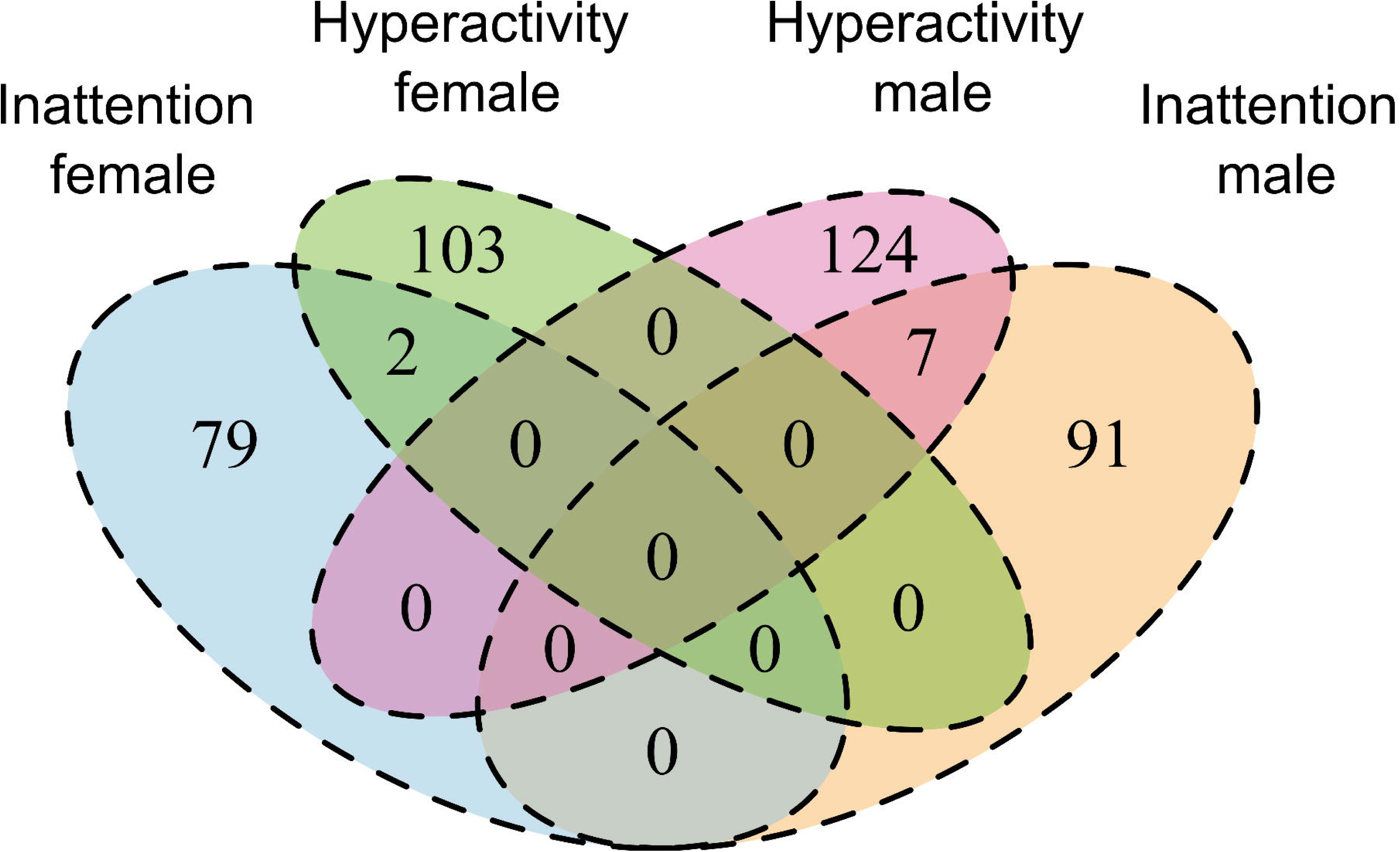
Venn diagram depicting the overlap of top CpGs (p-value < 0.0001) in cord blood associated with inattention or hyperactivity/impulsivity in female and male participants of the MoBa study.

## Discussion

Our sex-stratified epigenome-wide association meta-analysis of cord blood revealed that the DNA methylation landscape associated with childhood ADHD symptoms is largely sex-specific. The identification of sex-specific DMPs and DMRs, which became attenuated when female and male data were combined, highlights the importance of sex-informed analytical approaches. Furthermore, the epigenetic signatures of inattention and hyperactivity/impulsivity showed notable differences, suggesting distinct biological correlates for these clinical subtypes.

Exposure to an adverse intra-uterine environment, such as maternal obesity, hypertension, and medications, contributes to the risk of ADHD.^36^ The DNA methylation marks we identified in cord blood may reflect the biological embedding of *in utero* exposure to one or more adversities.^37^ The identified DMPs are unlikely to have resulted from the influence genetic variations (i.e., they do not have known *me*QTLs)^34^ suggesting they may reflect environmental or developmental effects. Nonetheless, a role for genetic influences is still possible given the recognized effect of gene-environment interaction in shaping the epigenome.^38^

Genes (*ARRB1*, *KDM5B*, and *PNPO*) annotated to the identified DMPs and DMRs have known roles in nervous system development and neurotransmission. For instance, *PNPO* is crucial for the synthesis of vitamin B6, a cofactor in the biosynthesis of monoamine neurotransmitters.^39,40^ Additionally, the vitamin B6 analogue (metadoxine) has been shown to attenuate symptoms of ADHD.^41^ Similarly, mutations in *KDM5B* have been implicated in developmental disorders, including ADHD and autism spectrum disorder.^35,42,43^ *ARRB1* encodes a protein, β-arrestin 1, which plays a key role in dampening intracellular β-adrenergic neuronal signaling and may also be involved in immune system regulation.^44^ While the tissue specificity of DNA methylation must be considered, the convergence of these annotated genes on plausible neurobiological pathways for ADHD supports the potential of these cord blood signatures as markers of early-life risk.

The molecular mechanisms for the sex differences in prevalence and clinical presentation of ADHD are not well understood.^8^ While a genome-wide association study has not found remarkable sex differences in genetic architecture,^45^ our findings demonstrate notable sex differences at the epigenetic level. The lack of overlap between the DNA methylation signatures in males and females indicates that the peripheral epigenetic profiles associated with ADHD symptoms differ at birth. The identification of a larger number of DNA methylation signatures in females, despite comparable sample sizes, is intriguing. More importantly, the female-specific cord blood DNA methylation signatures hold translational promise. They could form the basis for biomarkers to help identify at-risk females, thereby mitigating diagnostic disparities and enabling timely intervention.^3,4,46,47^

Our stratification by symptom domain also provides insight into the heterogeneity of ADHD. The contrasting DNA methylation signatures of inattention and hyperactivity/impulsivity enhance our understanding of their potentially distinct biological correlates and complement reported differences in genetic architecture.^48^ Such stratification is crucial for biological discovery and may eventually have relevance for patient stratification in clinical settings.^9,49^ While there was some overlap of DMRs across the symptom domains, the overall pattern suggests disparate methylation signatures, supporting the merit of parsing ADHD into its constituent subtypes.

A key methodological implication of our work is that sex-stratified analyses, despite reduced sample sizes, improve power for identifying associations in phenotypes with pronounced sex differences. The significant sex interaction effects we observed may explain the diminished discovery in our sex-differentiated meta-analyses, as effects present in only one sex are attenuated when groups are combined.^18,50^ As shown in our results, epigenome-wide significant DMPs in one sex typically showed no association in the other. Therefore, future epigenetic studies of ADHD and other disorders with sex differences should routinely employ sex-stratified analyses to advance discovery.

The following limitations must be considered when interpreting our findings. First, DNA methylation in cord blood may not directly reflect changes in the brain, limiting the ability to draw causal inferences about central nervous system pathophysiology. However, our goal was to discover early-life peripheral biomarkers of risk, for which cord blood is an accessible tissue. Second, the generalizability of our findings to diverse populations requires further investigation, though we adjusted for ancestry to ensure the inclusion of the minority of MoBa participants from diverse backgrounds. Key strengths include the relatively large sample size and the use of a dimensional measure of ADHD symptoms, which reduces sex biases reported in categorical ADHD diagnoses.^3,4^

In conclusion, the findings provide evidence for sex-specific and subtype-specific cord blood DNA methylation signatures of childhood ADHD. These indicate that peripheral epigenetic correlates of ADHD are evident at birth and show pronounced sex differences. The findings also support sex-stratified approaches to enhance epigenetic discoveries and suggest the potential for these early-life markers to improve early detection, particularly in females with symptoms of inattention. Future research is needed to replicate these findings and assess the predictive value of sex-specific polymethylation scores to advance the development of biomarkers for risk stratification.

## Data Availability

EWAS summary data will become freely available on Zenodo.org upon publication of the manuscript.

## Acknowledgments

The Norwegian Mother, Father and Child Cohort Study is supported by the Norwegian Ministry of Health and Care Services and the Ministry of Education and Research. This research is part of the HARVEST collaboration, supported by the Research Council of Norway (grant 229624). The Norwegian Centre for Mental Disorders Research (NORMENT) provided genotype data, funded by the Research Council of Norway (grant 223273), South East Norway Health Authorities, and Stiftelsen Kristian Gerhard Jebsen. The Center for Diabetes Research, the University of Bergen, provided genotype data funded by the ERC AdG project SELECTionPREDISPOSED, Stiftelsen Kristian Gerhard Jebsen, the Trond Mohn Foundation, the Research Council of Norway, the Novo Nordisk Foundation, the University of Bergen, and the Western Norway Health Authorities. We thank Dr. Stephanie London for providing feedback on a previous version of the manuscript. The authors would like to acknowledge support from the Research Council of Norway (RCN) (273291, 273446) and the European Economic Area and Norway Grants (EEA-RO-NO-2018-0573). This work was performed on the TSD (Tjeneste for Sensitive Data) facilities, owned by the University of Oslo, operated and developed by the TSD service group at the University of Oslo, IT-Department (USIT).

## Conflict of Interest

The authors have no conflicts of interest to declare.

## Availability of Data and Materials

EWAS summary statistics will become freely available on Zenodo.

## Notes

### Competing Interest Statement

The authors have declared no competing interest.

### Author Declarations

The Norwegian Data Protection Agency and the Regional Committees for Medical and Health Research Ethics approved the establishment of MoBa and the initial data collection. Participating mothers provided written informed consent. The MoBa study is regulated by the Norwegian Health Registry Act. The current study was approved by the Regional Committees for Medical and Health Research Ethics (REK 2018/1898).

### Summary of Updates

The manuscript results have been updated after re-analysis of data after removing duplicate individuals. Supplementary files include more detailed figures, and tables. The discussion section has been updated inline with the changes in the new results section.

## References

1. Ayano G, Demelash S, Gizachew Y, Tsegay L, Alati R. The global prevalence of attention deficit hyperactivity disorder in children and adolescents: An umbrella review of meta-analyses. J Affect Disord. Oct 15 2023;339:860–866. doi:10.1016/j.jad.2023.07.071

2. Cortese S, Song M, Farhat LC, et al. Incidence, prevalence, and global burden of ADHD from 1990 to 2019 across 204 countries: data, with critical re-analysis, from the Global Burden of Disease study. Mol Psychiatry. Nov 2023;28(11):4823–4830. doi:10.1038/s41380-023-02228-3

3. Mowlem FD, Rosenqvist MA, Martin J, Lichtenstein P, Asherson P, Larsson H. Sex differences in predicting ADHD clinical diagnosis and pharmacological treatment. Eur Child Adolesc Psychiatry. Apr 2019;28(4):481–489. doi:10.1007/s00787-018-1211-3

4. Martin J, Langley K, Cooper M, et al. Sex differences in attention-deficit hyperactivity disorder diagnosis and clinical care: a national study of population healthcare records in Wales. J Child Psychol Psychiatry. Dec 2024;65(12):1648–1658. doi:10.1111/jcpp.13987

5. Huang CL, Weng SF, Ho CH. Gender ratios of administrative prevalence and incidence of attention-deficit/hyperactivity disorder (ADHD) across the lifespan: A nationwide population-based study in Taiwan. Psychiatry Res. Oct 30 2016;244:382–7. doi:10.1016/j.psychres.2016.08.023

6. Taylor MJ, Martin J, Butwicka A, et al. A twin study of genetic and environmental contributions to attention-deficit/hyperactivity disorder over time. J Child Psychol Psychiatry. Nov 2023;64(11):1608–1616. doi:10.1111/jcpp.13854

7. Faraone SV, Bellgrove MA, Brikell I, et al. Attention-deficit/hyperactivity disorder. Nat Rev Dis Primers. Feb 22 2024;10(1):11. doi:10.1038/s41572-024-00495-0

8. Babinski DE. Sex Differences in ADHD: Review and Priorities for Future Research. Curr Psychiatry Rep. Apr 2024;26(4):151–156. doi:10.1007/s11920-024-01492-6

9. Cecil CAM, Nigg JT. Epigenetics and ADHD: Reflections on Current Knowledge, Research Priorities and Translational Potential. Mol Diagn Ther. Nov 2022;26(6):581–606. doi:10.1007/s40291-022-00609-y

10. Spiers H, Hannon E, Schalkwyk LC, et al. Methylomic trajectories across human fetal brain development. Genome Res. Mar 2015;25(3):338–52. doi:10.1101/gr.180273.114

11. Solomon O, Huen K, Yousefi P, et al. Meta-analysis of epigenome-wide association studies in newborns and children show widespread sex differences in blood DNA methylation. Mutat Res Rev Mutat Res. Jan-Jun 2022;789:108415. doi:10.1016/j.mrrev.2022.108415

12. Neumann A, Walton E, Alemany S, et al. Association between DNA methylation and ADHD symptoms from birth to school age: a prospective meta-analysis. Transl Psychiatry. Nov 12 2020;10(1):398. doi:10.1038/s41398-020-01058-z

13. Walton E, Pingault JB, Cecil CA, et al. Epigenetic profiling of ADHD symptoms trajectories: a prospective, methylome-wide study. Mol Psychiatry. Feb 2017;22(2):250–256. doi:10.1038/mp.2016.85

14. Meijer M, Klein M, Hannon E, et al. Genome-Wide DNA Methylation Patterns in Persistent Attention-Deficit/Hyperactivity Disorder and in Association With Impulsive and Callous Traits. Front Genet. 2020;11:16. doi:10.3389/fgene.2020.00016

15. Rovira P, Sanchez-Mora C, Pagerols M, et al. Epigenome-wide association study of attention-deficit/hyperactivity disorder in adults. Transl Psychiatry. Jun 19 2020;10(1):199. doi:10.1038/s41398-020-0860-4

16. van Dongen J, Zilhao NR, Sugden K, et al. Epigenome-wide Association Study of Attention-Deficit/Hyperactivity Disorder Symptoms in Adults. Biol Psychiatry. Oct 15 2019;86(8):599–607. doi:10.1016/j.biopsych.2019.02.016

17. Mooney MA, Ryabinin P, Wilmot B, Bhatt P, Mill J, Nigg JT. Large epigenome-wide association study of childhood ADHD identifies peripheral DNA methylation associated with disease and polygenic risk burden. Transl Psychiatry. Jan 21 2020;10(1):8. doi:10.1038/s41398-020-0710-4

18. Tesfaye M, Spindola LM, Stavrum AK, et al. Sex effects on DNA methylation affect discovery in epigenome-wide association study of schizophrenia. Mol Psychiatry. Aug 2024;29(8):2467–2477. doi:10.1038/s41380-024-02513-9

19. Magnus P, Birke C, Vejrup K, et al. Cohort Profile Update: The Norwegian Mother and Child Cohort Study (MoBa). Int J Epidemiol. Apr 2016;45(2):382–8. doi:10.1093/ije/dyw029

20. Haberg SE, Page CM, Lee Y, et al. DNA methylation in newborns conceived by assisted reproductive technology. Nat Commun. Apr 7 2022;13(1):1896. doi:10.1038/s41467-022-29540-w

21. Silva RR, Alpert M, Pouget E, et al. A rating scale for disruptive behavior disorders, based on the DSM-IV item pool. Psychiatr Q. Winter 2005;76(4):327–39. doi:10.1007/s11126-005-4966-x

22. Elliott HR, Tillin T, McArdle WL, et al. Differences in smoking associated DNA methylation patterns in South Asians and Europeans. Clin Epigenetics. Feb 3 2014;6(1):4. doi:10.1186/1868-7083-6-4

23. Zeilinger S, Kuhnel B, Klopp N, et al. Tobacco smoking leads to extensive genome-wide changes in DNA methylation. PLoS One. 2013;8(5):e63812. doi:10.1371/journal.pone.0063812

24. Houseman EA, Accomando WP, Koestler DC, et al. DNA methylation arrays as surrogate measures of cell mixture distribution. BMC Bioinformatics. May 8 2012;13:86. doi:10.1186/1471-2105-13-86

25. Ronningen KS, Paltiel L, Meltzer HM, et al. The biobank of the Norwegian Mother and Child Cohort Study: a resource for the next 100 years. Eur J Epidemiol. 2006;21(8):619–25. doi:10.1007/s10654-006-9041-x

26. Teschendorff AE, Marabita F, Lechner M, et al. A beta-mixture quantile normalization method for correcting probe design bias in Illumina Infinium 450 k DNA methylation data. Bioinformatics. Jan 15 2013;29(2):189–96. doi:10.1093/bioinformatics/bts680

27. Phipson B, Lee S, Majewski IJ, Alexander WS, Smyth GK. Robust Hyperparameter Estimation Protects against Hypervariable Genes and Improves Power to Detect Differential Expression. Ann Appl Stat. Jun 2016;10(2):946–963. doi:10.1214/16-AOAS920

28. Hoffler KD, Katrinli S, Halvorsen MW, et al. Optimizing genetic ancestry adjustment in DNA methylation studies: a comparative analysis of approaches. Epigenetics Chromatin. Oct 14 2025;18(1):69. doi:10.1186/s13072-025-00627-0

29. Mansell G, Gorrie-Stone TJ, Bao Y, et al. Guidance for DNA methylation studies: statistical insights from the Illumina EPIC array. BMC Genomics. May 14 2019;20(1):366. doi:10.1186/s12864-019-5761-7

30. Mallik S, Odom GJ, Gao Z, Gomez L, Chen X, Wang L. An evaluation of supervised methods for identifying differentially methylated regions in Illumina methylation arrays. Brief Bioinform. Nov 27 2019;20(6):2224–2235. doi:10.1093/bib/bby085

31. Pedersen BS, Schwartz DA, Yang IV, Kechris KJ. Comb-p: software for combining, analyzing, grouping and correcting spatially correlated P-values. Bioinformatics. Nov 15 2012;28(22):2986–8. doi:10.1093/bioinformatics/bts545

32. Cavalcante RG, Sartor MA. annotatr: genomic regions in context. Bioinformatics. Aug 1 2017;33(15):2381–2383. doi:10.1093/bioinformatics/btx183

33. Maksimovic J, Oshlack A, Phipson B. Gene set enrichment analysis for genome-wide DNA methylation data. Genome Biol. Jun 8 2021;22(1):173. doi:10.1186/s13059-021-02388-x

34. Gaunt TR, Shihab HA, Hemani G, et al. Systematic identification of genetic influences on methylation across the human life course. Genome Biol. Mar 31 2016;17:61. doi:10.1186/s13059-016-0926-z

35. Harrington J, Wheway G, Willaime-Morawek S, Gibson J, Walters ZS. Pathogenic KDM5B variants in the context of developmental disorders. Biochim Biophys Acta Gene Regul Mech. Jul 2022;1865(5):194848. doi:10.1016/j.bbagrm.2022.194848

36. Kim JH, Kim JY, Lee J, et al. Environmental risk factors, protective factors, and peripheral biomarkers for ADHD: an umbrella review. Lancet Psychiatry. Nov 2020;7(11):955–970. doi:10.1016/S2215-0366(20)30312-6

37. Ma Z, Wang Y, Quan Y, Wang Z, Liu Y, Ding Z. Maternal obesity alters methylation level of cytosine in CpG island for epigenetic inheritance in fetal umbilical cord blood. Hum Genomics. Aug 31 2022;16(1):34. doi:10.1186/s40246-022-00410-2

38. Czamara D, Eraslan G, Page CM, et al. Integrated analysis of environmental and genetic influences on cord blood DNA methylation in new-borns. Nat Commun. Jun 11 2019;10(1):2548. doi:10.1038/s41467-019-10461-0

39. Stover PJ, Field MS. Vitamin B-6. Adv Nutr. Jan 2015;6(1):132–3. doi:10.3945/an.113.005207

40. Ng J, Heales SJ, Kurian MA. Clinical features and pharmacotherapy of childhood monoamine neurotransmitter disorders. Paediatr Drugs. Aug 2014;16(4):275–91. doi:10.1007/s40272-014-0079-z

41. Manor I, Newcorn JH, Faraone SV, Adler LA. Efficacy of metadoxine extended release in patients with predominantly inattentive subtype attention-deficit/hyperactivity disorder. Postgrad Med. Jul 2013;125(4):181–90. doi:10.3810/pgm.2013.07.2689

42. Faundes V, Newman WG, Bernardini L, et al. Histone Lysine Methylases and Demethylases in the Landscape of Human Developmental Disorders. Am J Hum Genet. Jan 4 2018;102(1):175–187. doi:10.1016/j.ajhg.2017.11.013

43. Olfson E, Farhat LC, Liu W, et al. Rare de novo damaging DNA variants are enriched in attention-deficit/hyperactivity disorder and implicate risk genes. Nat Commun. Jul 12 2024;15(1):5870. doi:10.1038/s41467-024-50247-7

44. Lorton D, Bellinger DL. Molecular mechanisms underlying beta-adrenergic receptor-mediated cross-talk between sympathetic neurons and immune cells. Int J Mol Sci. Mar 11 2015;16(3):5635–65. doi:10.3390/ijms16035635

45. Martin J, Walters RK, Demontis D, et al. A Genetic Investigation of Sex Bias in the Prevalence of Attention-Deficit/Hyperactivity Disorder. Biol Psychiatry. Jun 15 2018;83(12):1044–1053. doi:10.1016/j.biopsych.2017.11.026

46. Hinshaw SP, Nguyen PT, O’Grady SM, Rosenthal EA. Annual Research Review: Attention-deficit/hyperactivity disorder in girls and women: underrepresentation, longitudinal processes, and key directions. J Child Psychol Psychiatry. Apr 2022;63(4):484–496. doi:10.1111/jcpp.13480

47. Novik TS, Hervas A, Ralston SJ, et al. Influence of gender on attention-deficit/hyperactivity disorder in Europe--ADORE. Eur Child Adolesc Psychiatry. Dec 2006;15 Suppl 1:I15–24. doi:10.1007/s00787-006-1003-z

48. Greven CU, Rijsdijk FV, Plomin R. A twin study of ADHD symptoms in early adolescence: hyperactivity-impulsivity and inattentiveness show substantial genetic overlap but also genetic specificity. J Abnorm Child Psychol. Feb 2011;39(2):265–75. doi:10.1007/s10802-010-9451-9

49. Yuan D, Zhang M, Huang Y, Wang X, Jiao J, Huang Y. Noradrenergic genes polymorphisms and response to methylphenidate in children with ADHD: A systematic review and meta-analysis. Medicine (Baltimore). Nov 19 2021;100(46):e27858. doi:10.1097/MD.0000000000027858

50. Zhou J, Xia Y, Li M, et al. A higher dysregulation burden of brain DNA methylation in female patients implicated in the sex bias of Schizophrenia. Mol Psychiatry. Nov 2023;28(11):4842–4852. doi:10.1038/s41380-023-02243-4

