## Supplementary Information for "Sex-Specific Cord Blood DNA Methylation Signatures for Childhood ADHD Symptoms"

[Figure S8: Manhattan plot of DNA methylation loci association with ADHD symptoms in sex-differentiated (female and male) meta-analysis of children participants of the Norwegian Mother, Father and Child Cohort (MoBa) study. Dotted line represents p-value = 9 ×10^-08^; Circled dots represent CpGs with a p-value < 1 ×10^-05^.	11](#_Toc215224014)

[Figure S9: Manhattan plot of DNA methylation loci association with inattention symptoms in sex-differentiated (female and male) meta-analysis of children participants of the Norwegian Mother, Father and Child Cohort (MoBa) study. Dotted line represents p-value = 9 ×10^-08^; Circled dots represent CpGs with a p-value < 1 ×10^-05^.	11](#_Toc215224015)

[Figure S10: Manhattan plot of DNA methylation loci association with inattention symptoms in EWAS meta-analysis of male children participants of the Norwegian Mother, Father and Child Cohort (MoBa) study. Dotted line represents p-value = 9 ×10^-08^; Circled dots represent CpGs with a p-value < 1 ×10^-05^.	12](#_Toc215224016)

[Figure S11: Manhattan plot of DNA methylation loci association with inattention symptoms in EWAS meta-analysis of female children participants of the Norwegian Mother, Father and Child Cohort (MoBa) study. Dotted line represents p-value = 9 ×10^-08^; Circled dots represent CpGs with a p-value < 1 ×10^-05^; Diamond shapes represent CpGs with a p-value < 9 ×10^-08^.	12](#_Toc215224017)

[Figure S12: Manhattan plot of DNA methylation loci association with hyperactivity/ impulsivity symptoms in sex-differentiated (female and male) meta-analysis of children participants of the Norwegian Mother, Father and Child Cohort (MoBa) study. Dotted line represents p-value = 9 ×10^-08^; Circled dots represent CpGs with a p-value < 1 ×10^-05^; Diamond shapes represent CpGs with a p-value < 9 ×10^-08^.	13](#_Toc215224018)

[Figure S13: Manhattan plot of DNA methylation loci association with hyperactivity/ impulsivity symptoms in EWAS meta-analysis of male children participants of the Norwegian Mother, Father and Child Cohort (MoBa) study. Dotted line represents p-value = 9 ×10^-08^; Circled dots represent CpGs with a p-value < 1 ×10^-05^; Diamond shapes represent CpGs with a p-value < 9 ×10^-08^.	13](#_Toc215224019)

[Figure S14: Manhattan plot of DNA methylation loci association with hyperactivity/ impulsivity symptoms in EWAS meta-analysis of female children participants of the Norwegian Mother, Father and Child Cohort (MoBa) study. Dotted line represents p-value = 9 ×10^-08^; Circled dots represent CpGs with a p-value < 1 ×10^-05^; Diamond shapes represent CpGs with a p-value < 9 ×10^-08^.	14](#_Toc215224020)

### **Figure S1:** Flow chart showing samples from the Norwegian Mother, Father and Child Cohort (MoBa) study included in EWAS of ADHD symptoms and the reasons for exclusion.

MoBa1 with methylation

data (n=1128)

With ADHD data

(n=747)

Unique samples

(n=735)

No ADHD data (n=381)

Duplicates (n=12)

Samples with complete

data (n=646)

No covariates (n=89)

MoBa2 with methylation

data (n=813)

With ADHD data

(n=474)

Unique samples

(n=474)

Duplicates (n=0)

No ADHD data (n=339)

Sample with complete data (n=334)

No covariates (n=140)

MoBa4 with methylation

data (n=1945)

With ADHD data (n=1007)

Unique samples

(n=981)

Duplicates (n=26)

No ADHD data (n=938)

Sample with complete

data (n=904)

No covariates (n=77)

With ADHD data (n=568)

Unique samples

(n=563)

MoBa8 with methylation

data (n=1238)

Duplicates (n=5)

No ADHD data (n=670)

Unique samples

(n=507)

No covariates (n=56)

### **Figure S2:** Quantile-quantile plots and lambda (λ) values of sex-stratified epigenome-wide association of cord blood DNA methylation and ADHD symptoms in females in four sub-studies of the Norwegian Mother, Father and Child Cohort (MoBa) study.

| 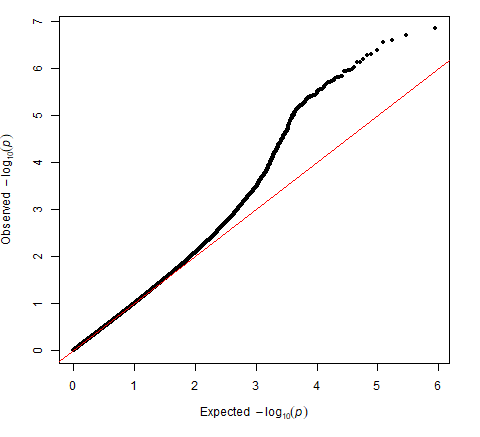   A. **MoBa1 (Met001) (λ = 0.98)** | 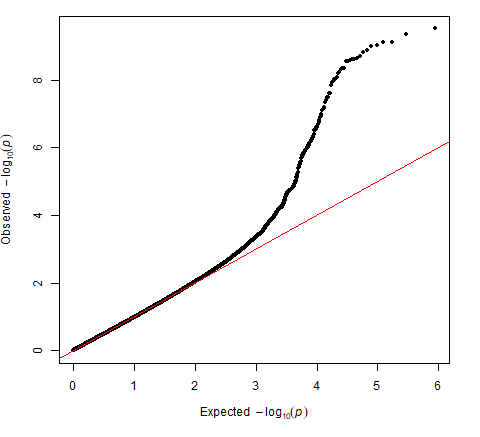  **B. MoBa2 (Met002) (λ = 0.96)** |
| --- | --- |
| 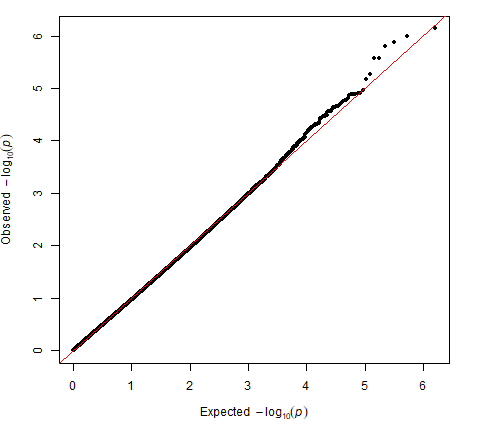   C. **MoBa4 (Met004) (λ = 0.95)** | 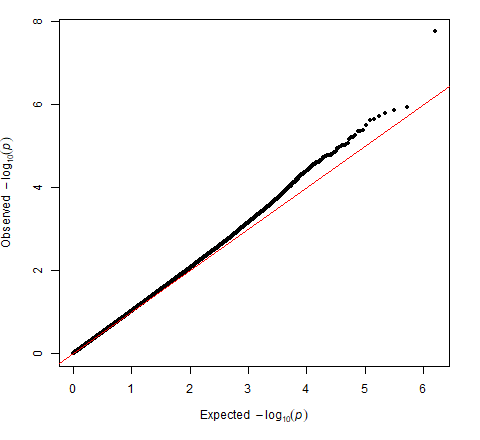  **D. MoBa8 (Met008) (λ = 1.03)** |

### **Figure S3:** Quantile-quantile plots and lambda (λ) values of sex-stratified epigenome-wide association of cord blood DNA methylation and ADHD symptoms in males in four sub-studies of the Norwegian Mother, Father and Child Cohort Study (MoBa).

| 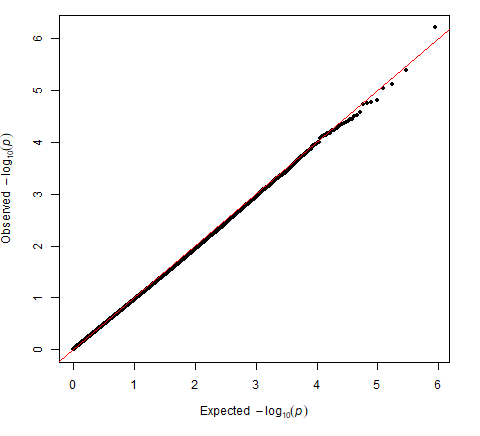   A. **MoBa1 (Met001) (λ = 0.95)** | 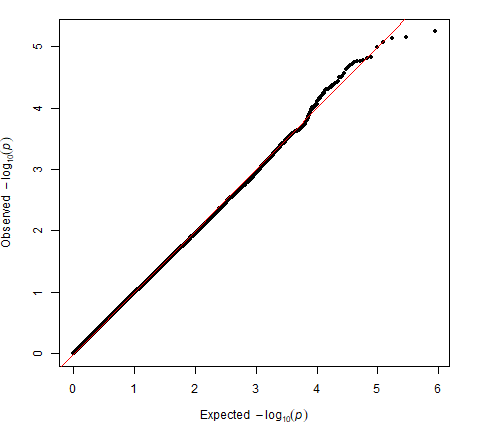  **B. MoBa2 (Met002) (λ = 0.98)** |
| --- | --- |
| 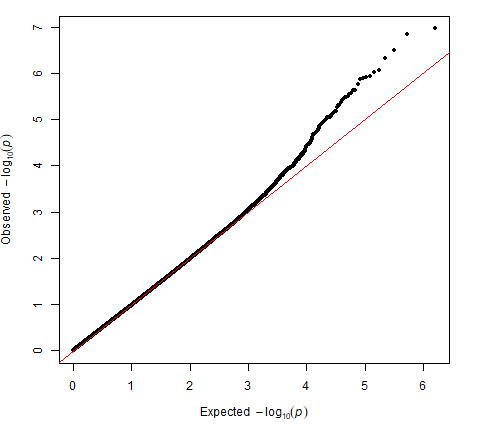   C. **MoBa4 (Met004) (λ = 0.97)** | 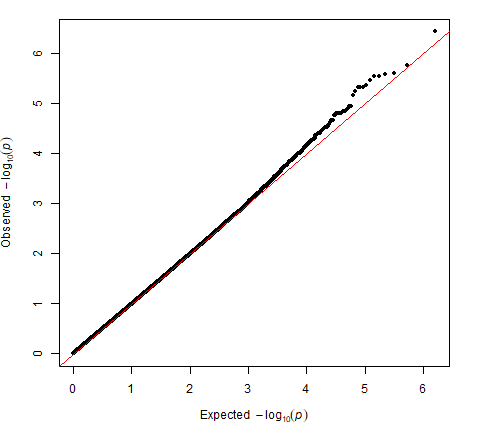  **D. MoBa8 (Met008) (λ = 0.99)** |

### **Figure S4:** Quantile-quantile plots and lambda (λ) values of sex-stratified epigenome-wide association of cord blood DNA methylation and inattention symptoms in females in four sub-studies of the Norwegian Mother, Father and Child Cohort (MoBa) study.

| 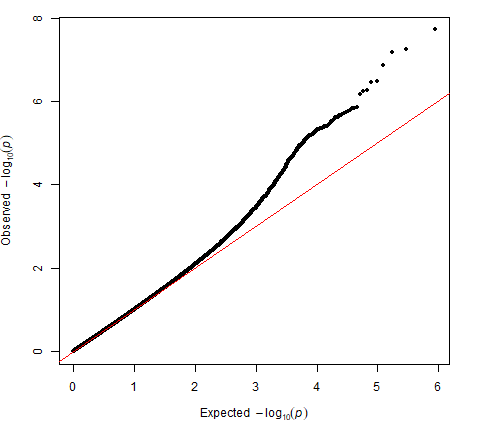   A. **MoBa1 (Met001) (λ = 0.99)** | 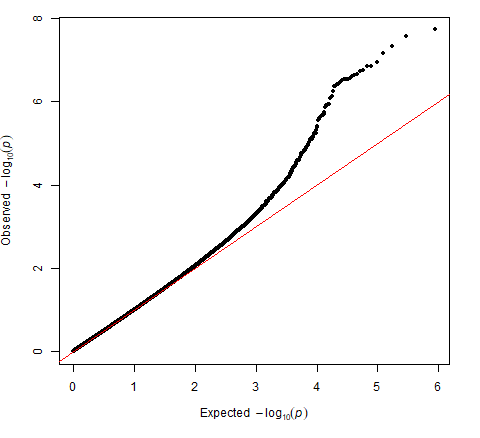  **B. MoBa2 (Met002) (λ = 0.99)** |
| --- | --- |
| 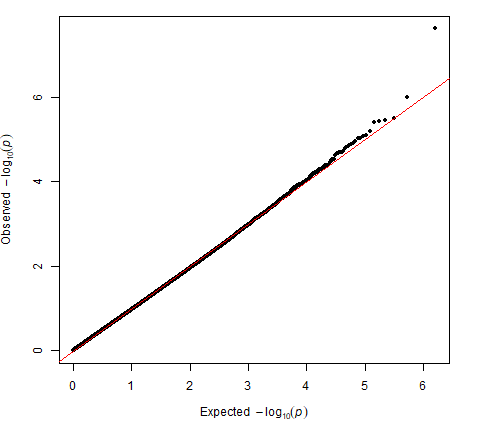   C. **MoBa4 (Met004) (λ = 0.95)** | 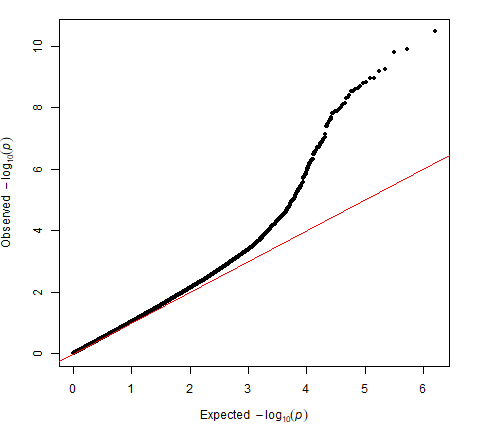  **D. MoBa8 (Met008) (λ = 1.03)** |

### **Figure S5:** Quantile-quantile plots and lambda (λ) values of sex-stratified epigenome-wide association of cord blood DNA methylation and inattention symptoms in males in four sub-studies of the Norwegian Mother, Father and Child Cohort (MoBa) study.

| 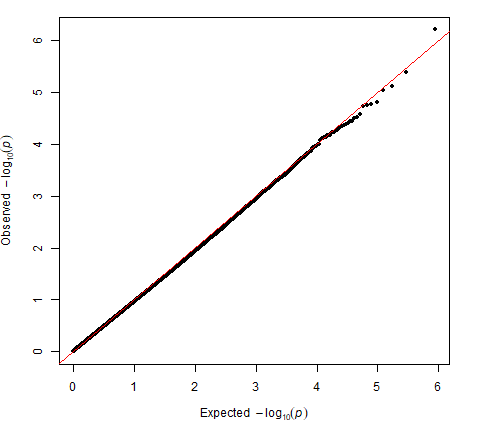   A. **MoBa1 (Met001) (λ = 0.95)** | 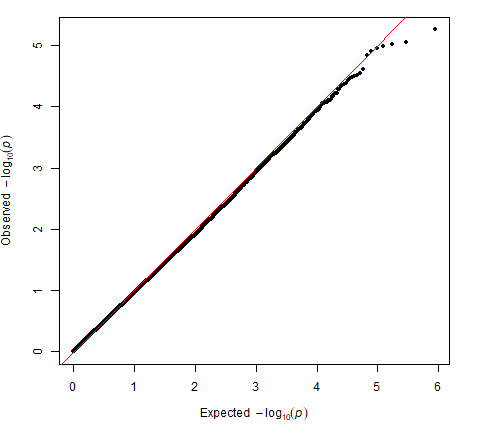  **B. MoBa2 (Met002) (λ = 0.95)** |
| --- | --- |
| 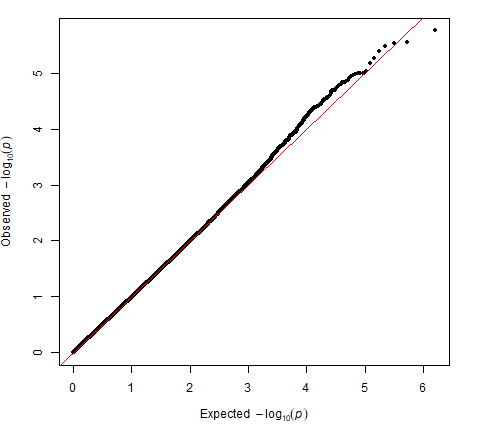   C. **MoBa4 (Met004) (λ = 0.99)** | 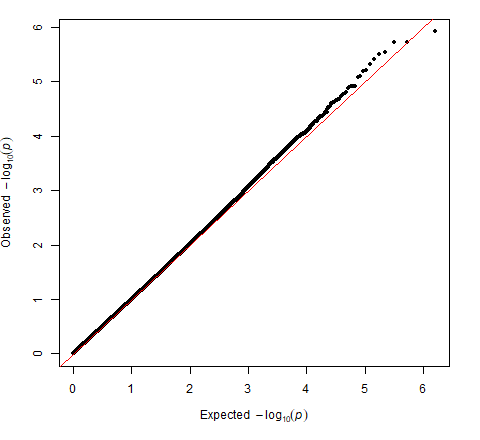  **D. MoBa8 (Met008) (λ = 0.99)** |

### **Figure S6:** Quantile-quantile plots and lambda (λ) values of sex-stratified epigenome-wide association of cord blood DNA methylation and hyperactivity/impulsivity symptoms in females in four sub-studies of the Norwegian Mother, Father and Child Cohort (MoBa) study.

| 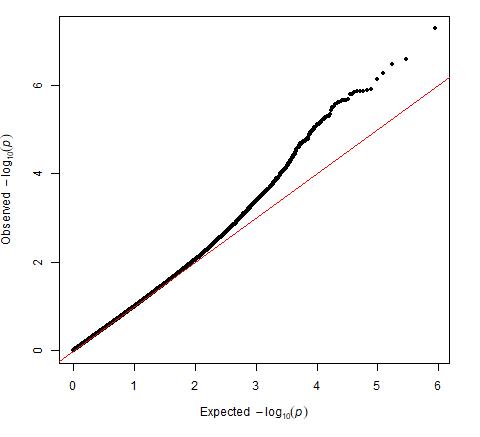   A. **MoBa1 (Met001) (λ = 0.98)** | 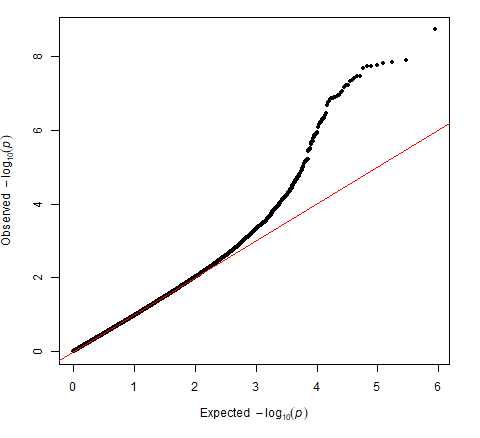  **B. MoBa2 (Met002) (λ = 0.95)** |
| --- | --- |
| 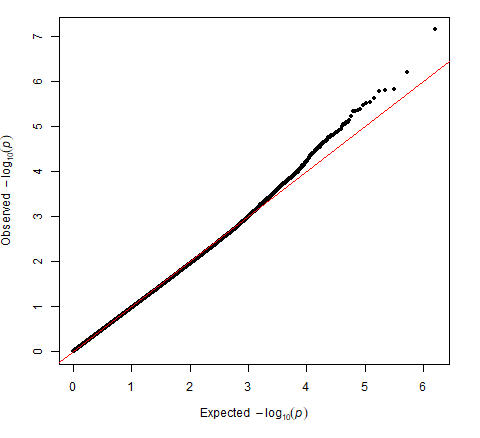   C. **MoBa4 (Met004) (λ = 0.96)** | 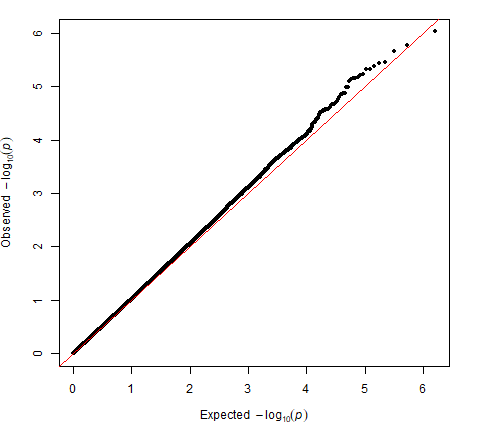  **D. MoBa8 (Met008) (λ = 1.02)** |

### **Figure S7:** Quantile-quantile plots and lambda (λ) values of sex-stratified epigenome-wide association of cord blood DNA methylation and hyperactivity/impulsivity symptoms in males in four sub-studies of the Norwegian Mother, Father and Child Cohort (MoBa) study.

| 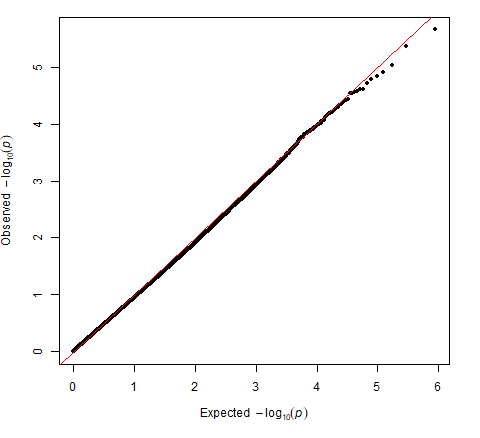   A. **MoBa1 (Met001) (λ = 0.93)** | 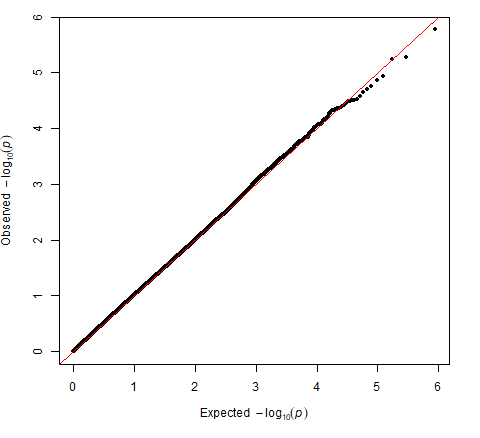  **B. MoBa2 (Met002) (λ = 1.03)** |
| --- | --- |
| 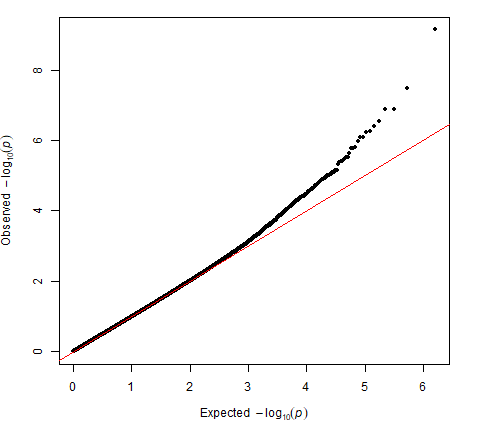   C. **MoBa4 (Met004) (λ = 0.97)** | 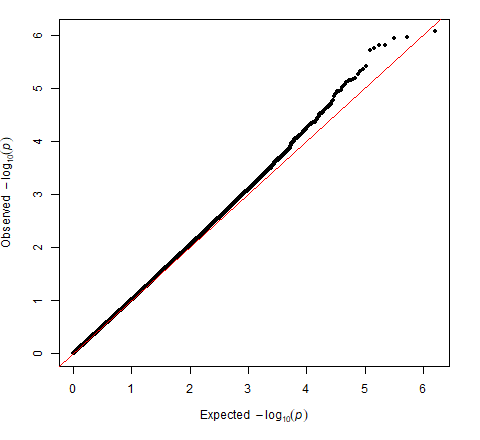  **D. MoBa8 (Met008) (λ = 1.01)** |

### **Figure S8:** Manhattan plot of DNA methylation loci association with ADHD symptoms in sex-differentiated (female and male) meta-analysis of children participants of the Norwegian Mother, Father and Child Cohort (MoBa) study. Dotted line represents p-value = 9 ×10^-08^; Circled dots represent CpGs with a p-value < 1 ×10^-05^.

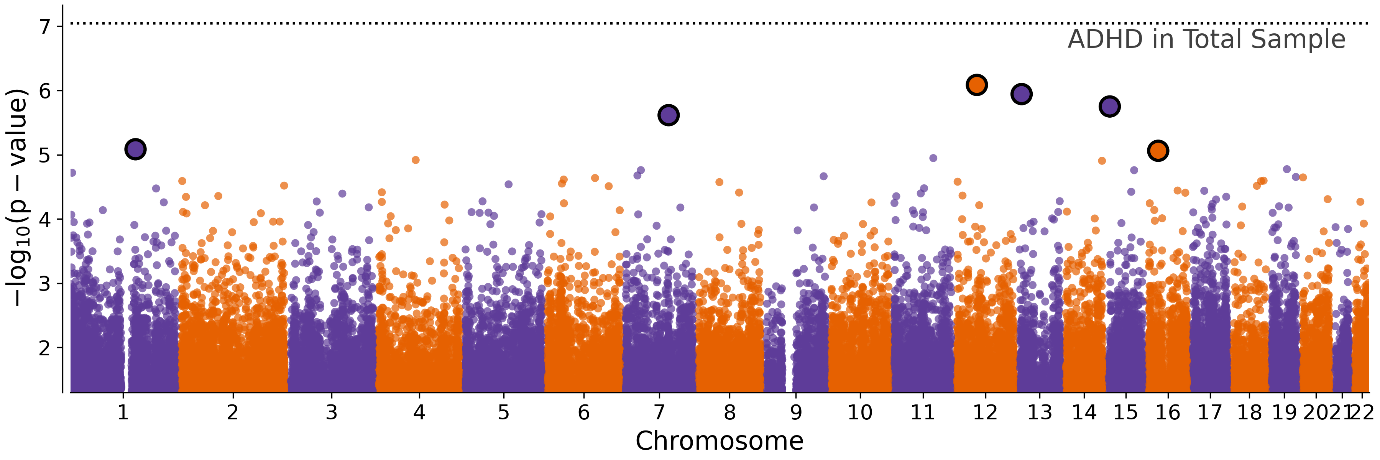

### **Figure S9:** Manhattan plot of DNA methylation loci association with inattention symptoms in sex-differentiated (female and male) meta-analysis of children participants of the Norwegian Mother, Father and Child Cohort (MoBa) study. Dotted line represents p-value = 9 ×10^-08^; Circled dots represent CpGs with a p-value < 1 ×10^-05^.

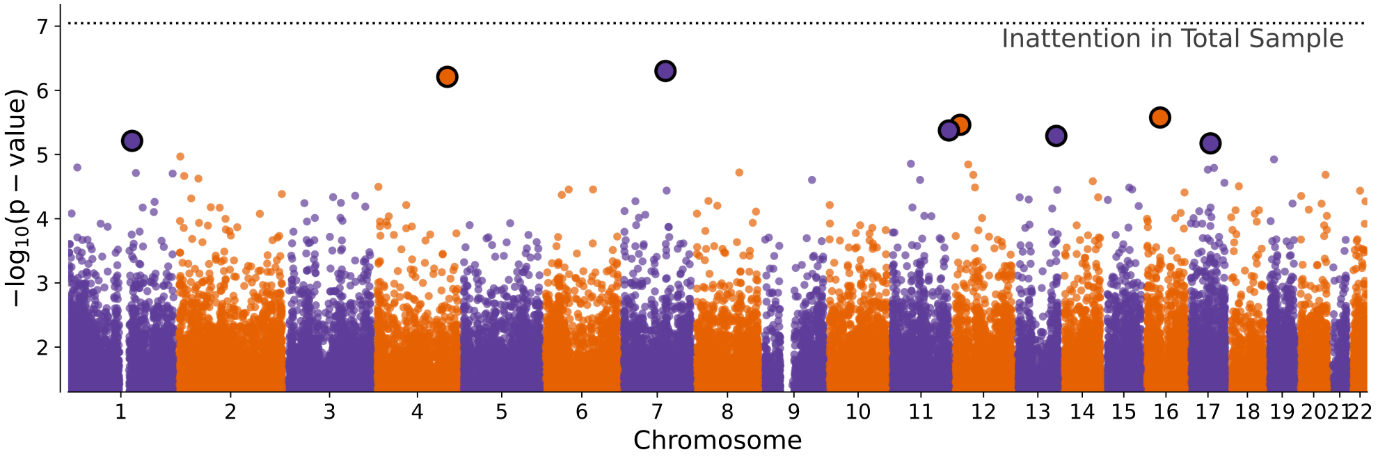

### **Figure S10:** Manhattan plot of DNA methylation loci association with inattention symptoms in EWAS meta-analysis of male children participants of the Norwegian Mother, Father and Child Cohort (MoBa) study. Dotted line represents p-value = 9 ×10^-08^; Circled dots represent CpGs with a p-value < 1 ×10^-05^.

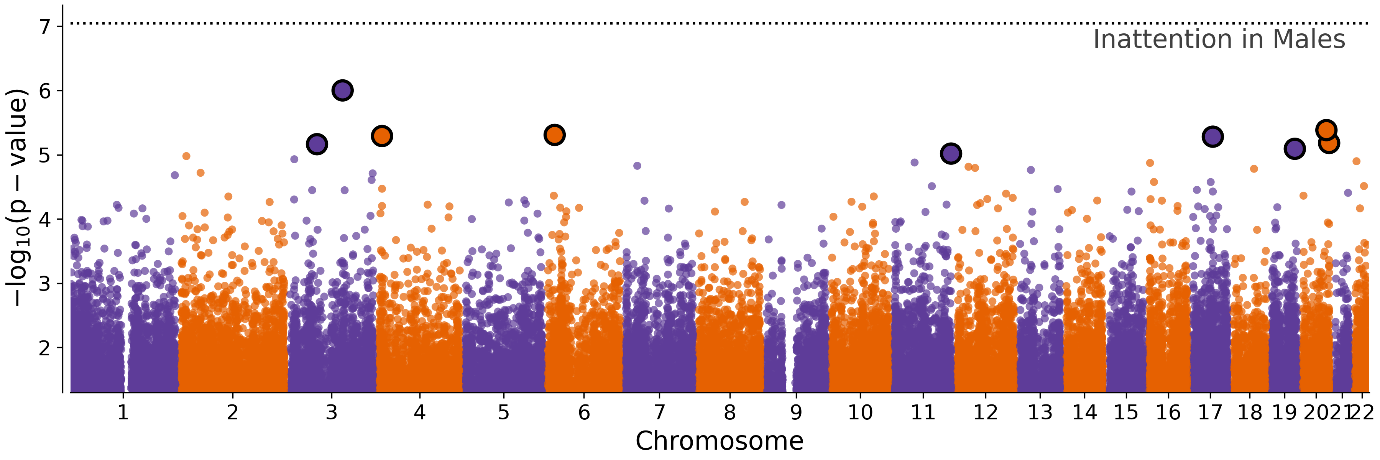

### **Figure S11:** Manhattan plot of DNA methylation loci association with inattention symptoms in EWAS meta-analysis of female children participants of the Norwegian Mother, Father and Child Cohort (MoBa) study. Dotted line represents p-value = 9 ×10^-08^; Circled dots represent CpGs with a p-value < 1 ×10^-05^; Diamond shapes represent CpGs with a p-value < 9 ×10^-08^.

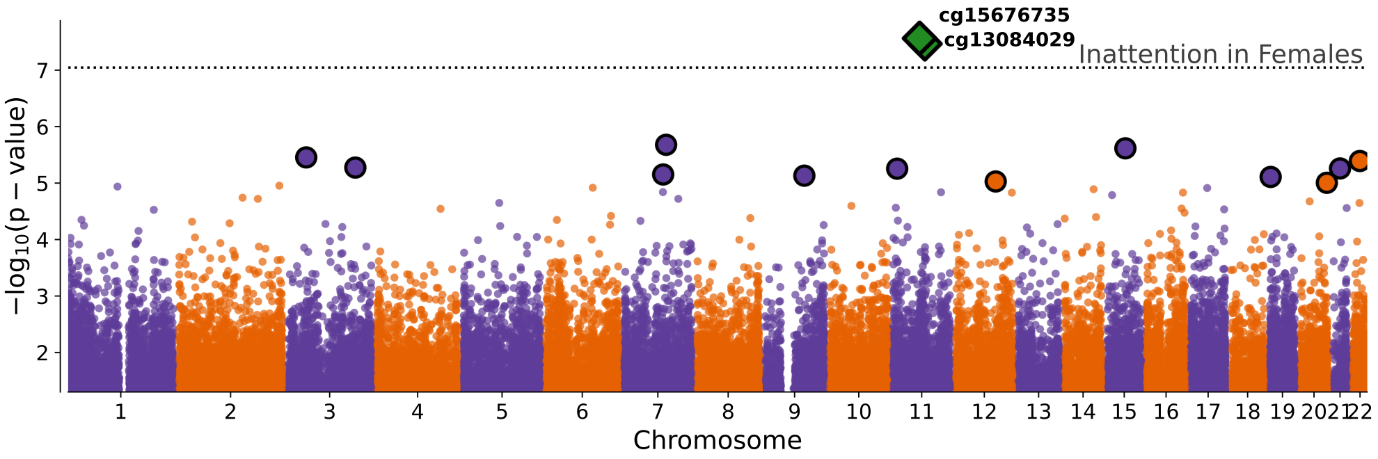

### **Figure S12:** Manhattan plot of DNA methylation loci association with hyperactivity/ impulsivity symptoms in sex-differentiated (female and male) meta-analysis of children participants of the Norwegian Mother, Father and Child Cohort (MoBa) study. Dotted line represents p-value = 9 ×10^-08^; Circled dots represent CpGs with a p-value < 1 ×10^-05^; Diamond shapes represent CpGs with a p-value < 9 ×10^-08^.

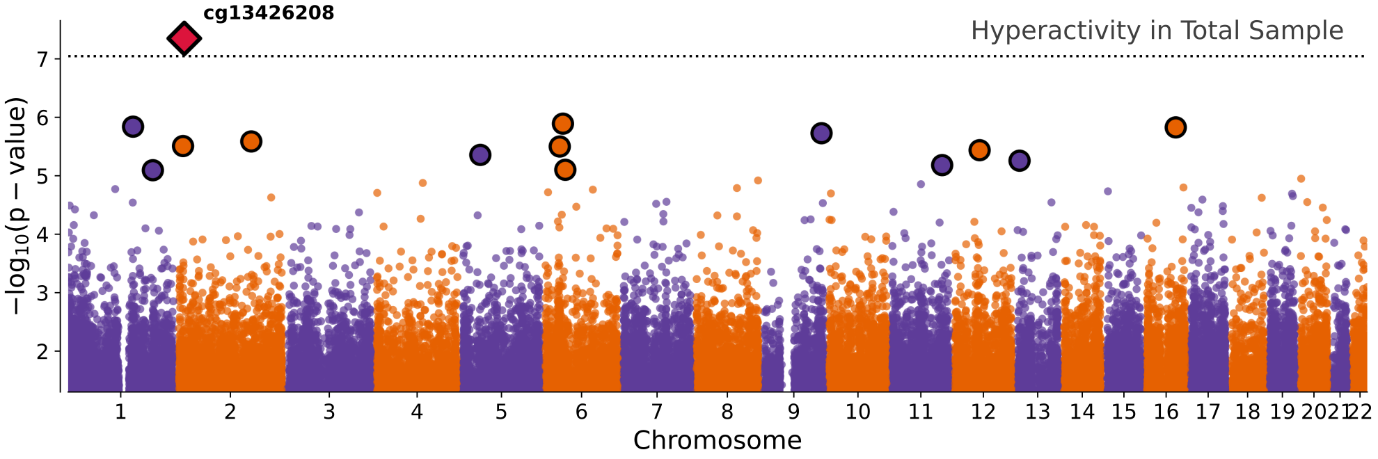

### **Figure S13:** Manhattan plot of DNA methylation loci association with hyperactivity/ impulsivity symptoms in EWAS meta-analysis of male children participants of the Norwegian Mother, Father and Child Cohort (MoBa) study. Dotted line represents p-value = 9 ×10^-08^; Circled dots represent CpGs with a p-value < 1 ×10^-05^; Diamond shapes represent CpGs with a p-value < 9 ×10^-08^.

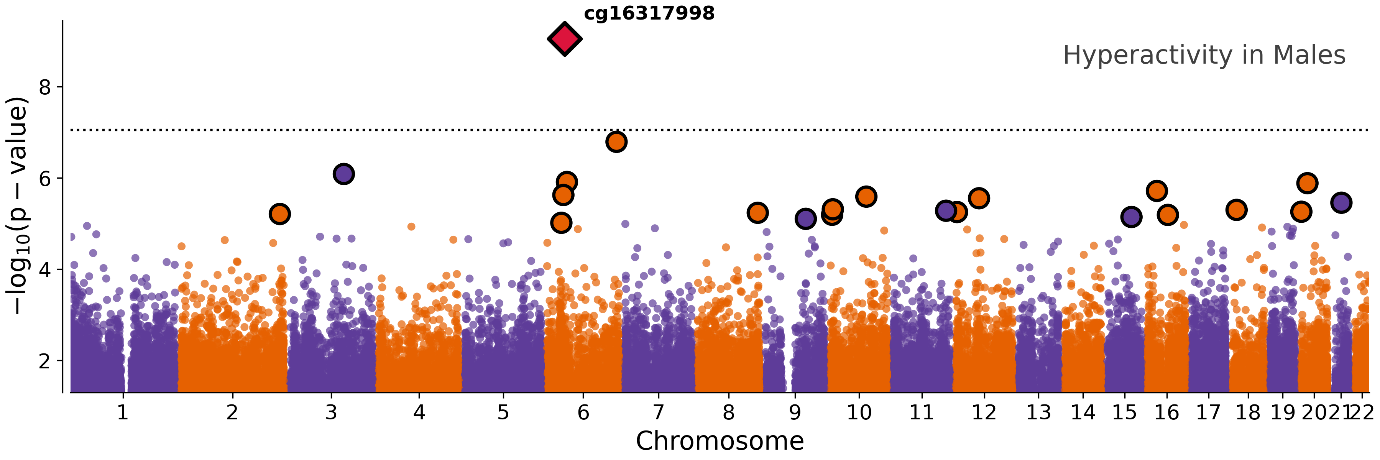

### **Figure S14:** Manhattan plot of DNA methylation loci association with hyperactivity/ impulsivity symptoms in EWAS meta-analysis of female children participants of the Norwegian Mother, Father and Child Cohort (MoBa) study. Dotted line represents p-value = 9 ×10^-08^; Circled dots represent CpGs with a p-value < 1 ×10^-05^; Diamond shapes represent CpGs with a p-value < 9 ×10^-08^.

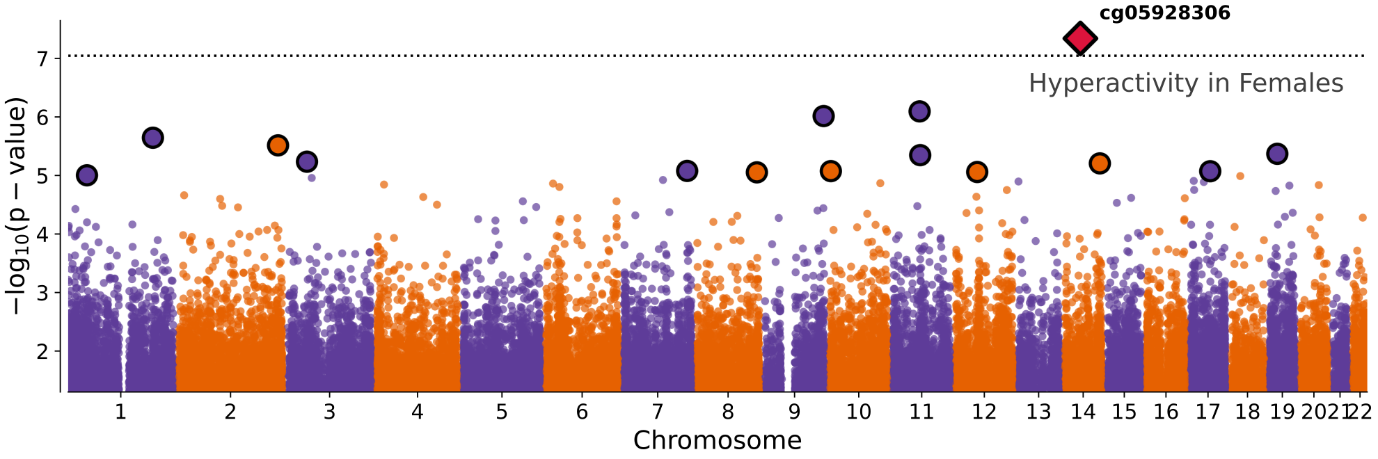
